# Analysis of time-to-positivity data in tuberculosis treatment studies: Identifying a new limit of quantification

**DOI:** 10.1101/2024.05.06.24306879

**Authors:** Suzanne M. Dufault, Geraint R. Davies, Elin M. Svensson, Derek J. Sloan, Andrew D. McCallum, Anu Patel, Pieter Van Brantegem, Paulo Denti, Patrick P. J. Phillips

## Abstract

The BACTEC Mycobacteria Growth Indicator Tube (MGIT) machine is the standard globally for detecting viable mycobacteria in patients’ sputum. Samples are observed for no longer than 42 days, at which point the sample is declared “negative” for tuberculosis (TB). This time to detection of bacterial growth, referred to as time-to-positivity (TTP), is increasingly of interest not solely as a diagnostic tool, but as a continuous biomarker wherein change in TTP over time can be used for comparing the bactericidal activity of different TB treatments. However, as a continuous measure, there are oddities in the distribution of TTP values observed, particularly at higher values. We explored whether there is evidence to suggest setting an upper limit of quantification (ULOQ_*M*_) lower than the diagnostic limit of detection (LOD) using data from several TB-PACTS randomized clinical trials and PanACEA MAMS-TB. Across all trials, less than 7.1% of all weekly samples returned TTP measurements between 25 and 42 days. Further, the relative absolute prediction error (%) was highest in this range. When modeling with ULOQ_*M*_ s of 25 and 30 days, the precision in estimation improved for 23 of 25 regimen-level slopes as compared to models using the diagnostic LOD while also improving the discrimination between regimens based on Bayesian posteriors. While TTP measurements between 25 days and the diagnostic LOD may be important for diagnostic purposes, TTP values in this range may not contribute meaningfully to its use as a quantitative measure, particularly when assessing treatment response, and may lead to under-powered clinical trials.

**Highlights:**

- The BACTEC Mycobacteria Growth Indicator Tube (MGIT) machine is the STAND, PaMZard globally for the detection and diagnosis of tuberculosis.
- As MGIT machine use becomes more ubiquitous, its time-to-positivity (TTP) measures are increasingly of interest as a continuous biomarker for evaluating bactericidal activity of TB treatment regimens.
- Using data from seven previously published trials, this work highlights the evidence for setting a limit of quantification for quantitative analyses that is below the diagnostic limit of detection. TTP values near the upper limit of detection appear to be noisier and sparser, with precision improving for estimation of 23 of 25 regimen-specific rates of change in TTP when analyzed with a lower limit of quantification.
- While TTP measurements between 25 days and the diagnostic LOD may be important for diagnostic purposes, TTP values in this range may not contribute meaningfully to its use as a quantitative measure, particularly when assessing early treatment response.

## 1 Introduction

*Mycobacterium tuberculosis* (Mtb), the primary pathogen responsible for tuberculosis (TB) disease, is a highly contagious, airborne bacterial species that has persisted across centuries and has been the leading cause of death by an infectious agent worldwide for decades, only outpaced in recent history by SARS-CoV-2 [1]. While mid-century treatment campaigns with novel antibiotic regimens provided optimism for the control of the disease, the emergence of multidrug-resistant Mtb strains and increased fatality rates in the co-occurring AIDS crisis heightened the priority for development of rapid diagnostics for TB. Augmenting the time-consuming microscopic examination of smears, which had served as the initial step in laboratory diagnosis with relatively low sensitivity, and the three-to-six week process of culturing and incubating samples on solid media, [2] the development of the BACTEC MGIT 960, a “fully automated, continuously monitoring, walk-away” system was revolutionary. The BACTEC MGIT machine incubates cultures in a liquid growth medium and includes a sensor that detects when oxygen is reduced by any aerobically metabolizing bacteria, substantially improving capacity, safety, and turnaround time for the detection of *M. tuberculosis* [3]. Manufacturers recommend samples are observed until a positive signal develops or for a maximum of 42 days, for the sake of diagnosis.

As MGIT tests have become more routine, TTP has become a useful biomarker in settings beyond diagnosis, including the evaluation of bactericidal activities of antibiotic regimens. Chigutsa et al. [4] proposed the first such model relating serial TTP measures to a patient’s time on treatment. TTP is a complex biomarker and modeling strategies must take into account the non-linearity of bactericidal activity, the right censoring induced by the manufacturer recommended diagnostic LOD of 42 days, and high participant variability in week-to-week measures of TTP. An example of both TTP’s rise in popularity as an endpoint and its complexity in modeling is observed in Study NC-005 (NCT02193776) of moxifloxacin (M), pretomanid (Pa), pyrazinamide (Z), and bedaquiline (B), (BPaMZ), which used the trajectory of weekly patient TTP measures as the primary outcome in a Phase II investigation of bactericidal activity of several new regimens. A Bayesian non-linear mixed effects regression model was used to accommodate the complexity in distribution and right censoring of the data at the diagnostic LOD of 42 days. Many other such models have since been proposed [4].

Despite tailoring models to account for many of the distributional oddities of TTP, we have observed across several studies, after contaminated samples have been excluded, fewer TTP values in the range of 25 to 42 than would be expected based on the distributional assumptions used to model TTP. While observations in this range may be important for individual diagnostic purposes, the TTP values in this range may not add value to a statistical model of regimen-level TTP trajectories, effectively suggesting there may be an upper limit of quantification for the sake of modeling, which we will refer to as ULOQ_*M*_. At worst, these observations may add noise, thereby reducing the ability of TTP modeling to measure treatment response and discriminate between regimens. We seek to test this hypothesis by examining the distributions of TTP data across several studies and assessing the evidence of a decreasing signal at higher values of TTP in replicate samples. We then explore the impact of different ULOQ_*M*_ thresholds on the estimation of model parameters, the precision in estimation, and the ability of the model to differentiate between regimens. By drawing across many case studies, we hope to avoid falling prey to overfitting, and aim to propose a ULOQ_*M*_ that will provide enhanced signal and precision when the objective is to model regimen-level treatment-response or exposure-response, necessary targets in identifying and characterizing promising regimens.

## 2 Materials and methods

### 2.1 Case Studies

We have gathered several case studies where TTP data has been collected at regular intervals, described briefly here and in Table 1.

**Table 1.**
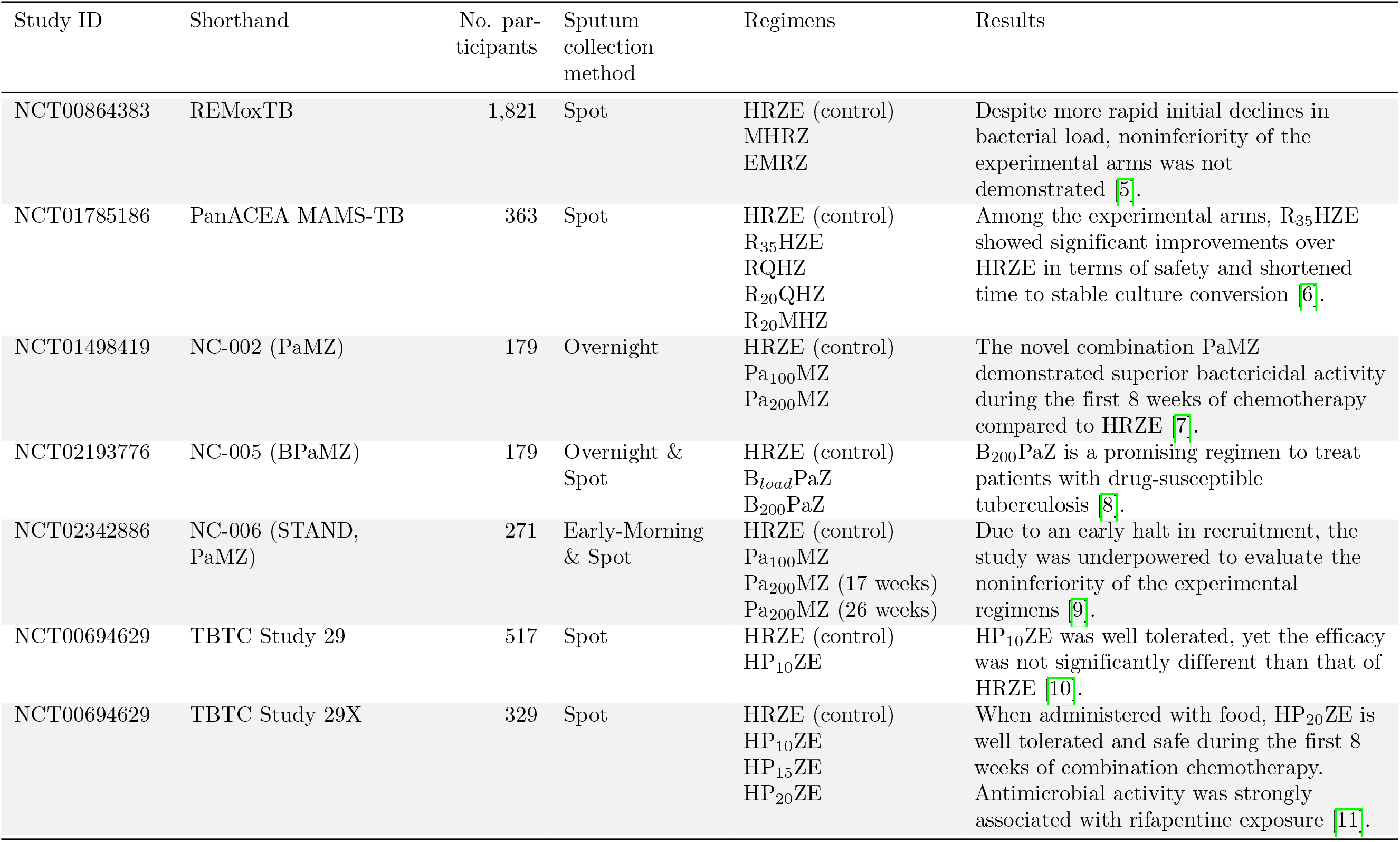
TBTC = Tuberculosis Trials Consortium. NC = TB Alliance New Combination. H = isoniazid, R = rifampicin at 10 mg/kg, Z = pyrazinamide, E = ethambutol, M = moxifloxacin, R_35_ = rifampicin at 35 mg/kg, Q = SQ109, R_20_ = rifampicin at 20 mg/kg, B = bedaquiline = bedaquiline at 400 mg/day for 14 days then 200 mg 3x/week, B_200_ = 200 mg/day, Pa_100_ = pretomanid at 100 mg, Pa_200_ = pretomanid at 2 P_10_ = rifapentine at 10 mg/kg, P_15_ = rifapentine at 15 mg/kg, P_20_ = rifapentine at 20 mg/kg.

#### 2.1.1 REMoxTB

The Rapid Evaluation of Moxifloxacin in Tuberculosis (REMoxTB) study (NCT00864383) was a large, randomized, placebo-controlled Phase III noninferiority study designed to evaluate if moxifloxacin (M) could replace either isoniazid (H) or ethambutol (E) in a four-month regimen for the treatment of TB [5]. Patients were randomized 1:1:1 to either the control arm (HRZE) or one of the novel four month regimens. Along with other biomarkers and endpoints, TTP was collected at baseline (pre-randomization), weekly for eight weeks post-randomization, and at monthly intervals until 26 weeks post-randomization. Sputum was decontaminated with acetylcysteine-sodium hydroxide [5]. The TTP data were initially used in secondary Kaplan-Meier analyses of time to culture-negative status based on measures taken from baseline until 78 weeks post-randomization. While improved bactericidal activity was observed in the novel regimens compared to the control, noninferiority based on the primary endpoint (unfavourable outcomes) was not demonstrated.

#### 2.1.2 PanACEA MAMS-TB

The PanACEA (Pan African Consortium for the Evaluation of Antituberculosis Antibiotics) multiple-arm, multiple-stage TB (PanACEA MAMS-TB) study (NCT01785186) was a large, randomized, open-label Phase II study designed to identify shorter, safer drug regimens for the treatment of TB [6]. Patients were randomized 2:1:1:1:1 to either the control arm (HRZE), or one of the novel four drug combinations consisting of rifampicin (R), isoniazid (H), pyrazinamide (Z), ethambutol (E), or SQ109 (Q) and/or moxifloxacin (M). Sputum was collected during clinic visits at a schedule of two days before start of treatment (pre-randomization), weekly for twelve weeks, and then at weeks 14, 17, 22, and 26 after treatment start. TTP played a pivotal role in this study, serving as the basis for the primary endpoint – time from treatment initiation to the first of two consecutive negative weekly sputum cultures – over the twelve-week window of observation. Among the experimental arms, only R_35_HZE showed significant improvements over the control in terms of safety and shortened time to stable culture conversion [6].

#### 2.1.3 TB-PACTS Datasets

TB-PACTS, a controlled access data platform with patient-level data from 26 TB trials (https://c-path.org/programs/tb-pacts/), is an invaluable resource for fueling TB research innovation. For this work, five trials, briefly described here, have been identified with regular, repeated TTP measurements. The trials were carried out by the TB Alliance New Combination (NC) and Tuberculosis Trial Consortium (TBTC) networks. All were open-label Phase II studies with the exception of NC-006 STAND, PaMZ (NCT02342886), which was an open-label Phase III study.

**NC-002 PaMZ (NCT01498419)** evaluated the safety, efficacy, and tolerability of moxifloxacin (M) plus pretomanid (Pa) plus pyrazinamide (Z) during the first 8 weeks of treatment of TB for drug-susceptible and multi-drug resistant TB. Overnight sputum samples were collected on the first three days of treatment then on days 7, 14, 21, 28, 35, 42, 49, and 56. Spot specimens were also collected at baseline, on days 1, 2, 3, and 7, and every second day until day 14 for the 14-day early bactericidal activity substudy. TTP was a secondary outcome, monitored in patients from baseline through eight weeks of follow-up. Both novel regimens had improved time to sputum culture negativity relative to HRZE, were well tolerated and had similar safety profiles. We make use of the 179 drug-sensitive participants whose TTP was available for evaluation in the secondary analysis. [7]

**NC-005 BPaMZ (NCT02193776)** aimed to determine the bactericidal activity of bedaquiline, moxifloxacin, pretomanid, and pyrazinamide regimens during eight weeks of treatment. TTP was the primary endpoint and was measured from baseline through eight weeks of treatment on 180 participants with drug-sensitive TB, with a unique sampling scheme. Two samples were collected per person, per week of follow-up: one collected at home overnight by the participant (“overnight”) and one collected at the site under the observation and guidance of the trial staff (“spot”). The overnight sputum sample was used in the primary analysis of the original trial. We will make use of the sputum results from the 179 drug-sensitive individuals [8].

**NC-006 Shortening Treatment by Advancing Novel Drugs (STAND, PaMZ)** trial (NCT02342886) aimed to assess the efficacy, safety and tolerability of four- and six-month durations of a novel regimen consisting of moxifloxacin (M), pretomanid (Pa), and pyrazinamide (Z). TTP was a secondary endpoint, with two samples collected per person, similar to the NC-005 sampling scheme, but replacing the overnight sample with one collected by the participant early in the morning (“early morning”). Due to an early halt in recruitment because of concerns regarding hepatotoxicity, the study was underpowered to evaluate the noninferiority of the experimental regimens [9]. We make use of the sputum results from the 271 drug-sensitive individuals [9].

**TBTC Study 29 (NCT00694629)** and its extension **TBTC Study 29X** examined the safety and efficacy of an experimental regimen comprised of rifapentine (P), isoniazid (H), pyrazinamide (Z), and ethambutol (E). TTP was measured every 2 weeks from baseline to eight weeks post-randomization and served as a secondary endpoint in order to explore its correlation with other clinical biomarkers and, more importantly, to culture conversion and treatment failure. We make use of the 517 (Study 29) and 329 (Study 29X) individuals whose TTP was available for evaluation [10, 11].

### 2.2 Visualizations

Our first objective was to visualize the trends in collected TTP data from baseline to eight weeks post-randomization. Though individual studies may collect TTP for longer durations, we have chosen eight weeks both to reflect the typical duration of Phase II studies and because the majority of samples are negative after this point, which means they can no longer contribute to a quantitative understanding of trend. Individual trajectory plots were created to provide insight into the noisiness of the raw data with smoothing splines capturing trends at the regimen-level. Alluvial plots were built to demonstrate trends across categorized TTP measures by week of observation. Histograms of the TTP results by week are also included in the Supplementary Material (Fig. S1).

### 2.3 Examination of Signal-to-Noise

Within the studies where replicate measures of TTP were available, we proposed a ULOQ_*M*_ through an investigation of signal-to-noise across the range of observable TTP values. We adapted the approach adopted by the International Council for Harmonisation of Technical Requirements for Pharmaceuticals for Human Use (ICH) Q14 guidelines, which is based on “the analyte concentration for which the relative prediction error is at most 10%.” [12] For analyses of TTP as a quantitative (rather than dichotomous diagnostic) measure, we offer the following parallel for this exploratory work: “the TTP limit for which the relative prediction error is at most 10%.”

### 2.4 Modeling

A linear model was used to relate the logarithm of measured TTP (i.e., *y* = log_10_(TTP)) for individual *i* = 1,. .., *N*_*j*_ in treatment group *j* = 1,. .., *J* at visit *k* = 1,. .., *T*_*ij*_ to the time since randomization *t* (Eq.

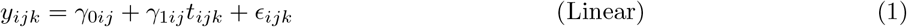

The objective of modeling the data was to examine the impact on the estimated posteriors for the parameter of interest (*γ*_1*j*_) when the TTP data were handled with different ULOQ_*M*_ as compared to the diagnostic LOD of 42 days. The impact of changing the ULOQ_*M*_ was measured by: 1) changes to the point estimates, 2) changes to the estimated precision for point estimates, and 3) changes in the posterior probabilities that the relative slope for a treatment group *γ*_1*j*_ as compared to the control is greater than or equal to some threshold, *τ* (i.e., Pr(*γ*_1*j*_*/γ*_1,HRZE_ *2: τ*)).

We use Bayesian estimation with weakly informative priors on the parameters (Supplemental Material) to fit these models. Estimation is performed with the “brms” package and visualized with “bayesplot” and “ggplot2” packages in R. All analysis code is available at a public GitHub repository maintained by the first author (https://github.com/sdufault15/tb-lod-ttp). Further details regarding model fit and assumptions can be found in the Supplemental Material.

## 3 Results

### 3.1 Visualizations

Using REMoxTB as an example, trends in the trajectories observed in TTP data are visualized in Fig 1. Fig 1A demonstrates the regimen-level trends, as fit by a smoothing spline. The two novel regimens (MHRZ and EMRZ) appear indistinguishable from each other, but both appear to have a faster rate of increasing TTP than the control regimen (HRZE). Fig 1B shows the individual-level trajectories, faceted by regimen. Individual trajectories have high variation from week to week. These trajectories also tend to increase over time; few individuals are observed to start at high or low TTP and remain fixed at those levels. The same visualization for the other datasets can be found in Figure S4.

**Fig 1.**
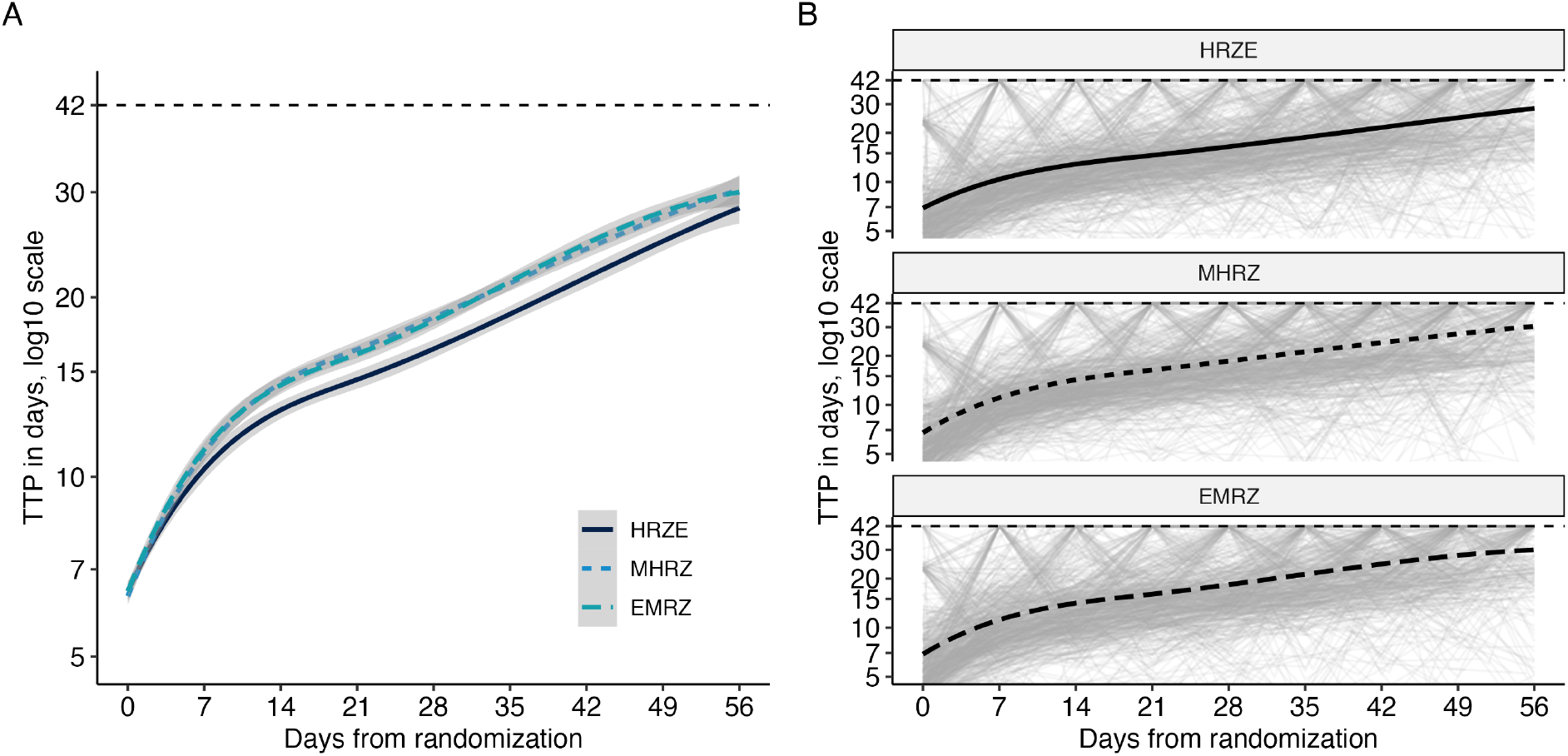
Observed time-to-positivity trajectories in REMoxTB. Any observations at or above the diagnostic limit of detection (42 days) are recorded as 42 days. A: Regimen-level trends in TTP (lines) and estimated STAND, PaMZard errors (ribbons) as fit by smoothing splines. B: Individual TTP trajectories (light gray) and regimen-level smoothing spline (black).

The noisiness of the individual-level trajectories is evident in Fig 1, but it is difficult to distinguish the paucity of samples returning TTP observations between 25 and 42 days. To directly examine this, Fig 2 displays the categorized distribution of the weekly TTP sample results. Two observations arise as expected: at baseline, bacterial growth is detected in nearly all samples in under 25 days, and, by the end of eight weeks on treatment, most no longer detect bacterial growth (i.e., observed TTP *2:* 42 days). Perhaps unexpectedly, the majority of samples seem to jump directly from detectable at less than 25 days to undetectable (TTP *2:* 42 days). In REMoxTB and PanACEA MAMS-TB, the studies with the largest number of samples, only 3.53% (520 of 14,734 samples) and 7.05% (218 of 3,092 samples), respectively, return a sputum TTP between 25 and 42 days. The values for the rest of the studies are included in Table S1.

**Fig 2.**
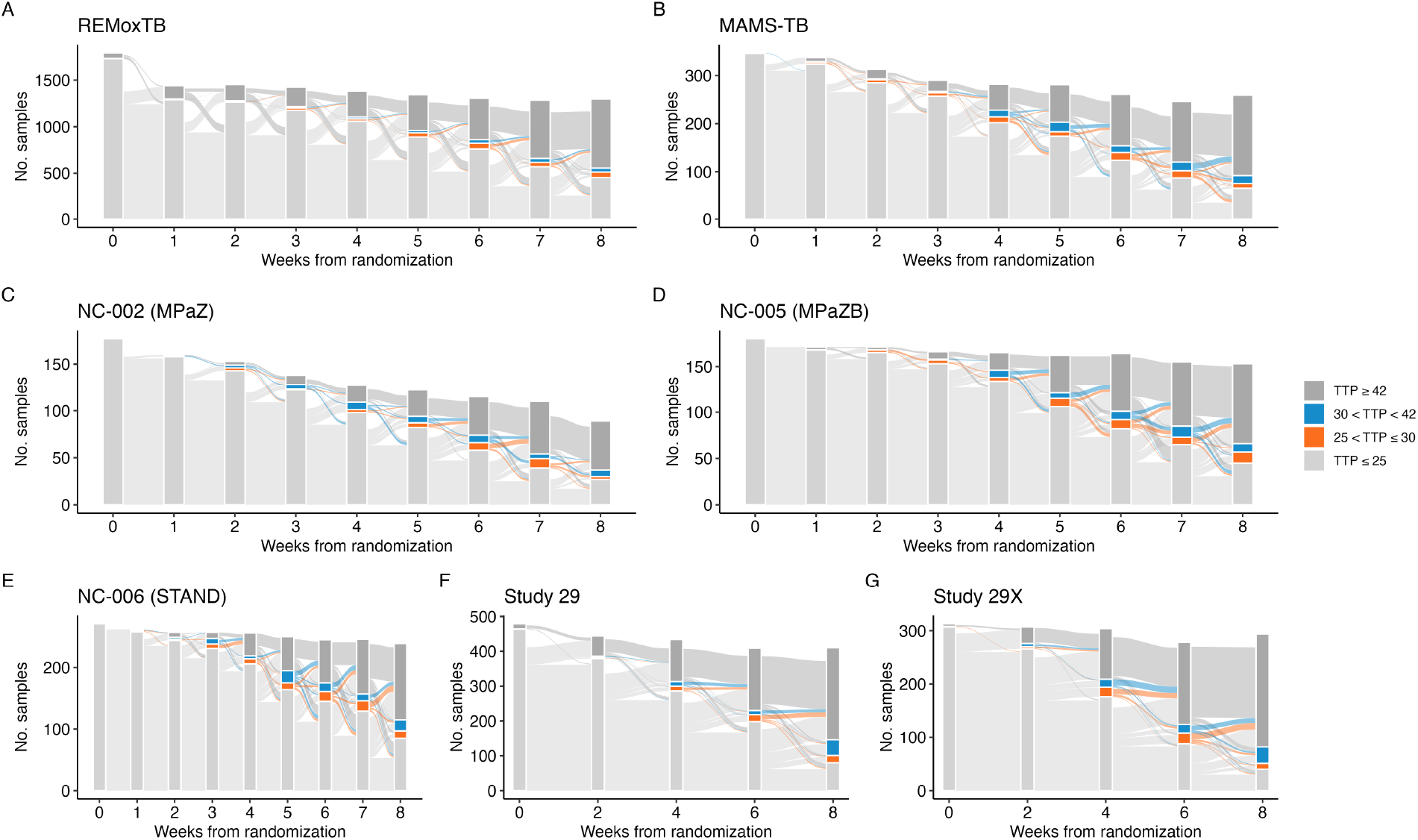
The flow of individuals’ weekly time-to-positivity (TTP) samples from measurements of ≤ 25 days, between 25 and 30 days, between 30 and 42 days, and above the diagnostic LOD (≥ 42 days) for A) REMox-TB, B) PanACEA MAMS-TB, C) NC-002 (PaMZ), D) NC-005 (BPaMZ), E) NC-006 (STAND, PaMZ), F) Study 29, and G) Study 29X.

### 3.2 Examination of Signal-to-Noise

Replicate data were available throughout the study period for Study NC-002 (PaMZ). The variation in correspondence of replicate measures can be seen in Fig. 3A where the replicates are plotted against each other. There are 1,003 replicated observations, none of which are replicate sample pairs that both returned observations above the diagnostic LOD. The correlation between all replicates (negative observations included and set to ‘42’) in NC-002 (PaMZ) is 77%, and 82.7% when restricted solely to the observations within the diagnostic LOD (negative observations excluded).

**Fig 3.**
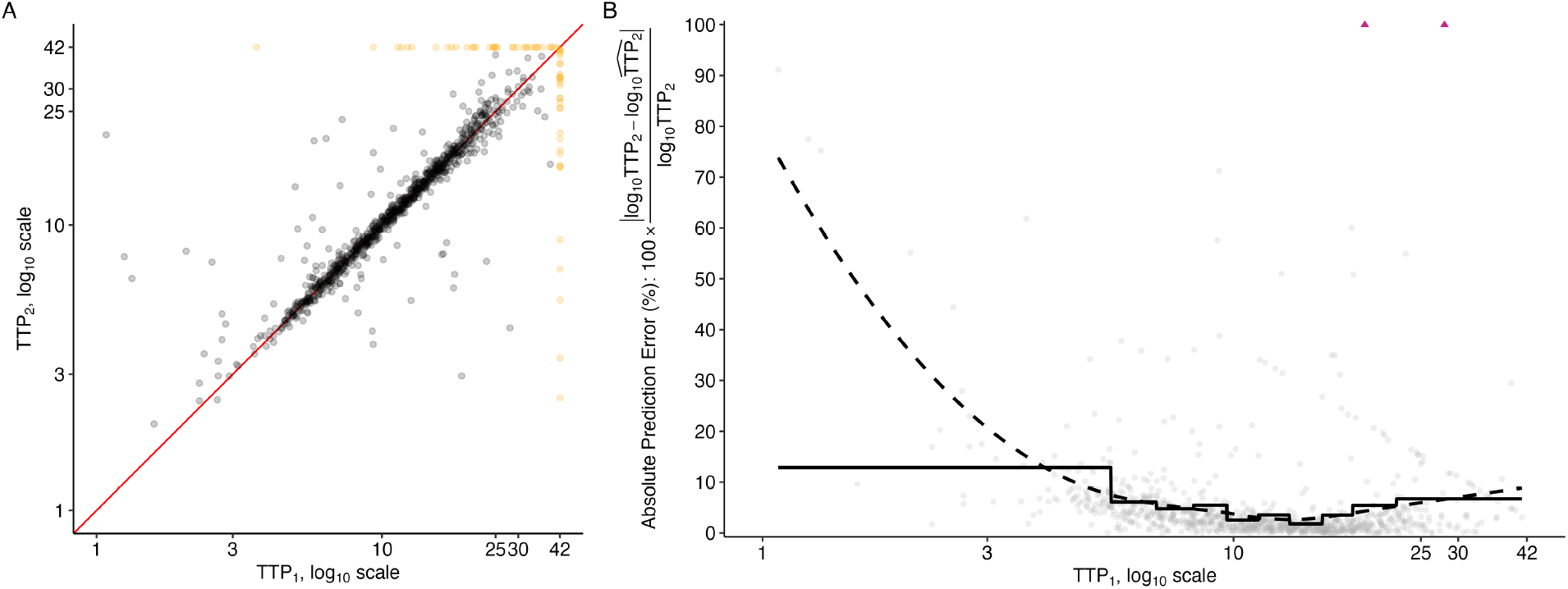
**A)** Replicate TTP observations from the NC-002 (PaMZ) study plotted against each other on the log_10_ scale. A red line indicates where perfect replication would lie. Points are marked in black (•) if below the diagnostic LOD and yellow 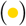 if above the diagnostic LOD. **B)** Absolute prediction error (%) when using one observation from each replicate pair (TTP_1_) to predict the second observation from each replicate pair (TTP_2_), using 5-fold cross-validation to train and test a simple linear prediction model. Data used for prediction is restricted to only those observations below the diagnostic limit of detection. The solid line denotes the average absolute prediction error (%) within deciles of TTP_1_ observations. The dashed line denotes a LOESS fit. Some predictions had an absolute prediction error (%) greater than 100% and are marked by triangles 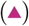.

**Fig 4.**
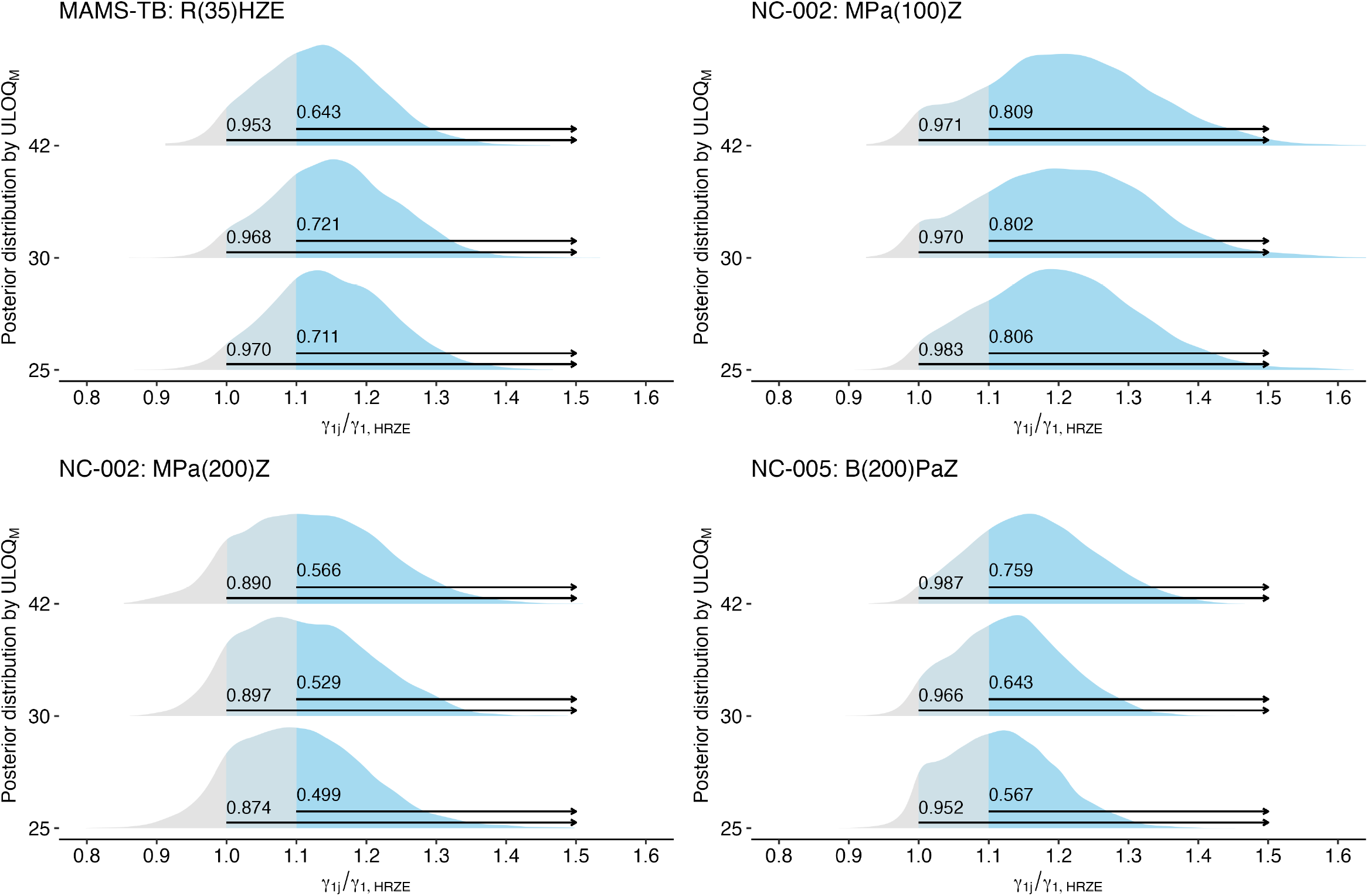
Among regimens with improved bactericidal activity over HRZE, the posterior distributions for the relative comparison of a regimen’s slope (*γ*_1*j*_) against the estimated slope on HRZE (*γ*_1,HRZE_), where a value of 1 indicates equal slopes and values > 1 suggest the regimen has greater bactericidal activity than HRZE. The estimated “confidence” that a regimen has any improvement in bactericidal activity over HRZE (Pr(*γ*_1*j*_*/γ*_1,HRZE_ > 1)) as well as the “confidence” that a regimen has more than 10% improvement in bactericidal activity over HRZE (Pr(*γ*_1*j*_*/γ*_1,HRZE_ > 1.1)) is indicated for each regimen at each ULOQ_*M*_.

When taking a closer look at the “signal-to-noise” available across the range of observable TTP, we see that over a substantial portion of the range of TTP values, the average prediction error is less than 20% when a simple linear model is used to predict one other observation from the replicate pair (log_10_TTP_2_) based on the other observation from the replicate pair (log_10_ TTP_1_). In both the LOESS-smoothed (Fig. 3B, dashed line) and interval-averaged estimates within deciles of the observed TTP_1_ values (Fig. 3B, solid line), the average absolute prediction error (%) takes a U-shape, with the lowest error corresponding to TTP values between 3 and 18 and rising for TTP values outside of this range.

As is evident from the solid line reflecting decile-averaged estimates of absolute prediction error (%) in Fig. 3, fewer than 10% of observations are in the upper observable range of TTP (i.e., values between 19 and 42). This makes determination of a single ULOQ_*M*_ essentially infeasible within this range. We therefore move forward with two proposed ULOQ_*M*_ s from this range: 25 and 30 days. Because so few samples return TTP between 25 and 42 days, there is a negligible effect in terms of available sample size when considering the various ULOQ_*M*_ s evaluated at each week of observation post-randomization, even when compared to those available under the diagnostic LOD (Fig S5).

### 3.3 Models

We move forward with the model in Eq. 1 applied to each of the datasets under the diagnostic LOD and the proposed ULOQ_*M*_ s. For 23 out of 25 regimens, there is an improvement in estimator precision when a lower ULOQ_*M*_ than the diagnostic LOD is applied. However, there is also a compression of the slope point estimates towards the null (Supplemental Material, Table S2 and Figure S2). For example, in the REMoxTB data, the HRZE has an estimated slope of 0.095 log_10_(TTP) per week since randomization (95% HCI: 0.090, 0.100) when the diagnostic LOD is applied. When a ULOQ_*M*_ = 25 is applied instead, the estimated slope decreases by 9.5% to 0.086 log_10_(TTP) per week since randomization yet the precision improves substantially resulting in a 20% decrease in the estimated credible interval width (95% HCI: 0.082, 0.090). Similar results can be seen for the other regimens and datasets in the Supplemental Material (Table S2 and Figure S2).

While the improvement in precision induced by the use of a lower ULOQ_*M*_ is a welcome result, the shift in point estimates towards the null means that such improved precision may not translate to improved differentiation in regimens’ bactericidal activity. To examine this directly, we examine the posterior probability that the relative slope for a treatment group *γ*_1*j*_ as compared to the control *γ*_1,HRZE_ is greater than or equal to some threshold, *τ* (i.e., Pr(*γ*_1*j*_*/γ*_1,HRZE_ *2: τ*)). First, we examine four regimens that were determined to have improved bactericidal activity in the clinical trial case studies (Table 1) to determine whether a change in the ULOQ_*M*_ may have improved the ability to differentiate these regimens from the control (HRZE). In the Bayesian linear models applied, three out of four regimens would have an improved estimated posterior probability (e.g., “confidence”) of greater early bactericidal activity (Pr(*γ*_1*j*_*/ γ*_1,HRZE_ *2:* 1)) if the ULOQ_*M*_ had been lower than the diagnostic LOD. In PanACEA MAMS-TB, a 25-day limit would have increased confidence of any improvement in early bactericidal activity from 95.3% to 97.0%. In NC-002, a 30-day limit would have made very little difference in the “confidence” of improvement in early bactericidal activity for MPa_100_Z (97.1% v. 97.0%), but would have slightly improved “confidence” for MPa_200_Z (89.0% v. 89.7%). For B_200_*PaZ* in NC-005 (PaMZ), a change in the ULOQ_*M*_ would have decreased confidence.

We also want to ensure that change in the ULOQ_*M*_ would not induce false confidence for regimens that were determined to be equivalent or worse than HRZE in terms of early bactericidal activity. To this end, we examined the posteriors associated with regimens from the PanACEA MAMS-TB case study that were determined to not have improved bactericidal activity relative to HRZE and found that the changes in “confidence” associated with changes in ULOQ_*M*_ would not have introduced a false positive result (Fig. S6).

## 4 Discussion

TTP is an increasingly utilized intermediate biomarker for the rapid evaluation of bactericidal activity of TB therapies. Across a series of case studies, we have demonstrated how few samples return TTP values between 25 and 42 days. While values in this range may be useful for diagnosis, the quantitative signal appears to be less reliable. Setting a lower ULOQ_*M*_ may improve precision and the ability to differentiate between novel regimens and the STAND, PaMZard of care, HRZE. We propose that in analyses where TTP is used as a continuous, quantitative measure, a ULOQ_*M*_ of 25 or 30 days is appropriate.

This is the first work, to our knowledge, to make the case of decreasing the ULOQ for modeling TTP. Other work has described the properties of TTP before, but primarily with regards to its suitability as an alternative to counting solid medium bacterial colony-forming units (CFUs), the predominant diagnostic and modeling biomarker used before the development of TTP.

The advantages of decreasing the limit of quantification include improved precision in 23 of 25 regimen-level slopes from the linear models applied across the case studies. Improving precision directly increases power and strengthens our ability to identify meaningful differences (tests of equivalence) or similarities (tests of noninferiority) when they are present. Operationally, a practical benefit is the ability to be adaptive earlier in trial settings when TTP models are used to assess regimens for futility at interim analyses. Observing samples for 25 rather than 42 days saves two weeks in terms of decision-making capacity, which means patients can be diverted away from regimens lacking evidence of effect and more quickly assigned to regimens that are demonstrating promise at early stages of clinical trials. This faster turnaround of results is increasingly important in the era of adaptive trial designs, where GO/NO-GO decisions are being made during interim analyses [13]; such a limit change may have a tangible impact on the efficiency with which modern trial designs can be implemented in the study of TB therapeutics. It is important to note that we are explicitly not advocating for the lowering of the limit of detection for diagnostic purposes.

There are disadvantages in setting a lower ULOQ_*M*_. It is hardly comfortable to recommend “throwing out” data. However, we have hoped to demonstrate that the data above the proposed limits is proportionally small and disproportionately noisy. While we cannot be certain that we are not trading bias for precision, the case studies have demonstrated that the changes in point estimates are substantially less than the decrease in variance.

Further research into the reasons behind the noisiness of TTP values above 25 days is warranted, and is beyond the scope of this paper. One possibility may be that the machinery itself is not well-calibrated for quantitative results in this range. For example, if the resources available in the MGIT tube decrease as the period of observation lengthens, the Mtb present may not grow exponentially. Another possibility concerns the time at which the sample was collected and its impact on the quantity, quality, and activity of the Mtb present. For instance, many of the TTP values above 25 days arise at later points in treatment and, therefore, may generally have a scarcity of Mtb present relative to samples earlier in treatment. The limited quantity of Mtb present in these samples may play a role in the noisiness in several ways. First, it simply may not withstand the necessary processing and dilution protocols, which would further explain the poor replicability observed here. Second, it may be more impacted by decontamination and sterilization procedures. Complete sterilization of other competitors without killing Mtb is likely not possible, but the consequences may not be visible when Mtb is abundant and capable of outgrowing competitors by orders of magnitude. As for the quality and activity of the Mtb, it may be possible that samples taken later in treatment are either more prone to contamination, and therefore more prone to being excluded from analyses such as these, or result in more contamination given a more dormant Mtb population. Unlike samples taken earlier in treatment, the Mtb produced in sputum later in treatment may be less active and take longer to grow. In the meantime, this provides a window of opportunity for other populations to establish, resulting in more contaminated samples.

It is also worth noting that the current treatment of TTP as a right-censored continuous variable is not the only approach that may be useful. When treated as a time-to-event variable, the limit of quantification and general challenges around right-censoring are less problematic. Such approaches have been demonstrated in semi-mechanistic models [14, 15]. However, semi-mechanistic models tend to have many parameters and are often unstable. Another option may be to consider a different error structure for the TTP in this upper range, perhaps implementing a power function or other method that would increase the uncertainty in this range. Work has been done in this area, but the distributional assumptions are often too complicated or uncertain for estimation purposes.

Further, it is apparent that TTP does not only appear to have an upper limit of quantification problem. Previous research has observed issues with left-censoring of TTP, perhaps due to the “bacterial lag phase induced by the sodium hydroxide-based decontamination procedure before MGIT inoculation, which could delay the onset of metabolic activity independent of the actual number of bacteria inoculated” [16]. We also observe this here in Figure S1.

## 5 Conclusion

The diagnostic limit of detection (LOD) for TTP may not be an appropriate upper limit of quantification (ULOQ_*M*_) when TTP is used as a continuous measure, particularly for the purposes of modeling and decision-making regarding regimen performance. TTP observations above 25 days appear to be rare and disproportionately noisy. While we cannot be certain that by applying a ULOQ_*M*_ that is less than the LOD we are not trading bias for precision, the case studies have demonstrated that any introduction of potential estimator bias is offset by gains in estimator precision and measurement signal.

## Data Availability

All datasets (with the exception of PanACEA MAMS-TB) analyzed during the current study are available in the TB-PACTS repository (https://c-path.org/programs/tb-pacts/). The MAMS-TB data can be requested from PanACEA executive group, reachable at: Postbus PanACEA secretariat.

## 8 Acknowledgments

We thank the PanACEA consortium for making the MAMS-TB data available to this project. The PanACEA consortium was funded by European and Developing Countries Clinical Trials partnership (grants IP.2007.32011.011, IP.2007.32011.012, and IP.2007.32011.013) and the German Ministry for Education and Research (grant 01KA0901).

## References

1. Global Tuberculosis Report 2022. Geneva: World Health Organization, 2022.

2. Tenover FC, Crawford JT, Huebner RE, Geiter LJ, Horsburgh CR, Good GC. The resurgence of tuberculosis: is your laboratory ready?. J Clin Microbiol. 1993 Apr;33(4):767–70.

3. Tortoli E, Cichero P, Piersimoni C, Simonetti MT, Gesu G, Nista D. Use of BACTEC MGIT 960 for recovery of mycobacteria from clinical specimens: multicenter study J Clin Microbiol. 1999 Nov;37(11):3578–82.

4. Chigutsa E, Patel K, Denti P, Visser M, Maartens G, Kirkpatrick CM, McIlleron H, Karlsson, MO. A Time-to-Event Pharmacodynamic Model Describing Treatment Response in Patients with Pulmonary Tuberculosis Using Days to Positivity in Automated Liquid Mycobacterial Culture. Antimicrob Agents Chemother. 2013 Feb;57(2):789–95.

5. Gillespie SH, Crook AM, McHugh TD, Mendel CM, Meredith SK, Murray SR, Pappas F, Phillips PPJ, Nunn AJ, REMoxTB Consortium Four-month moxifloxacin-based regimens for drug-sensitive tuberculosis New Engl J Med. 2014; 371(17):1577–1587.

6. Boeree MJ, Heinrich N, Aarnoutse R, Diacon AH, Dawson R, Rehal S, Kibiki GS, Churchyard G, Sanne I, Ntinginya, NE and others. High-dose rifampicin, moxifloxacin, and SQ109 for treating tuberculosis: a multi-arm, multi-stage randomised controlled trial The Lancet Infectious Diseases. 2017; 17(1):39–49.

7. Dawson R, Diacon AH, Everitt D, van Niekerk C, Donald PR, Burger DA, Schall R, Spigelman M, Conradie A, Eisenach K, and others. Efficiency and safety of the combination of moxifloxacin, pretomanid (PA-824), and pyrazinamide during the first 8 weeks of antituberculosis treatment: a phase 2b, open-label, partly randomised trial in patients with drug-susceptible or drug-resistant pulmonary tuberculosis The Lancet. 2015; 385(9979):1738–47.

8. Tweed CD, Dawson R, Burger DA, Conradie A, Crook AM, Mendel CM, Conradie F, Diacon AH, Ntinginya NE, Everitt DE and others. Bedaquiline, moxifloxacin, pretomanid, and pyrazinamide during the first 8 weeks of treatment of patients with drug-susceptible or drug-resistant pulmonary tuberculosis: a multicentre, open-label, partially randomised, phase 2b trial The Lancet Respiratory Medicine. 2019; 7(12):1048–1058.

9. Tweed, CD and Wills, GH and Crook, AM and Amukoye, E and Balanag, V and Ban, AYL and Bateson, ALC and Betteridge, MC and Brumskine, W and Caoili, J and others A partially randomised trial of pretomanid, moxifloxacin and pyrazinamide for pulmonary TB The International Journal of Tuberculosis and Lung Disease. 2021;25(4):305–14

10. Dorman SE, Goldberg S, Stout JE, Muzanyi G, Johnson JL, Weiner M, Bozeman L, Heilig CM, Feng P, Moro R and others. Substitution of rifapentine for rifampin during intensive phase treatment of pulmonary tuberculosis: study 29 of the tuberculosis trials consortium The Journal of infectious diseases. 2012;206(7)1030–40.

11. Dorman SE, Savic RM, Goldberg S, Stout JE, Schluger N, Muzanyi G, Johnson JL, Nahid P, Hecker EJ, Heilig CM and others. Daily rifapentine for treatment of pulmonary tuberculosis. A randomized, dose-ranging trial American journal of respiratory and critical care medicine. 2015;191(3)333–43.

12. Gegenschatz SA, Chiappini FA, Teglia CM, de la Peña AM, Goicoechea HC. Binding the gap between experiments, statistics, and method comparison: A tutorial for computing limits of detection and quantification in univariate calibration for complex samples. Analytica Chimica Acta. 2022 May 29;1209:339342.

13. Dufault SM, Crook AM, Rolfe K, Phillips PPJ. A flexible multi-metric Bayesian framework for decisionmaking in Phase II multi-arm multi-stage studies. Statistics in Medicine. 2024 Feb; 43(3):501–513.

14. Koele SE, Phillips PPJ, Upton CM, van Ingen J, Simonsson USH, Diacon AH, Aarnoutse RE, Svensson EM. Early bactericidal activity studies for pulmonary tuberculosis: A systematic review of methodological aspects. Int J Antimicrob Agents. 2023 May;61(5):106775.

15. Gausi K, Ignatius EH, Sun X, Kim S, Moran L, Wiesner L, von Groote-Bidlingmaier F, Hafner R, Donahue K, Vanker N, Rosenkranz SL, Swindells S, Diacon AH, Nuermberger EL, Dooley KE, Denti P. A Semimechanistic Model of the Bactericidal Activity of High-Dose Isoniazid against Multidrug-Resistant Tuberculosis: Results from a Randomized Clinical Trial. Am J Respir Crit Care Med. 2021 Dec;204(11):1327–35.

16. Diacon AH, Maritz JS, Venter A, van Helden PD, Dawson R, Donald PR. Time to liquid culture positivity can substitute for colony counting on agar plates in early bactericidal activity studies of antituberculosis agents. Clin Microbiol Infect. 2012;18(7):711–7.

17. Kruschke JK. Bayesian analysis reporting guidelines. Nature Human Behavior. 2021;5(10):1282–91.

18. Stan Development Team Stan User’s Guide - Truncation and Censoring. https://mc-stan.org/docs/stan-users-guide/truncation-censoring.html#censored-data Accessed: 2024-03-27.

