## Supplemental file for "Analysis of time-to-positivity data in tuberculosis treatment studies: Identifying a new limit of quantification"

### Supplemental Material

#### S1.1 Proportion of all TTP samples in [25,42] range

**Table S1.** The proportion of all sputum samples with TTP values in the range [25,42) for sputum samples collected from baseline to eight weeks post-randomization.

| Trial | No. Samples | Proportion of Samples |
|---|---|---|
| REMoxTB | 520 | 0.035 |
| PanACEA MAMS-TB | 218 | 0.071 |
| NC-002 (PaMZ) | 140 | 0.046 |
| NC-005 (BPaMZ) | 174 | 0.048 |
| NC-006 (STAND, PaMZ) | 362 | 0.079 |
| Study 29 | 137 | 0.052 |
| Study 29X | 128 | 0.071 |

### S1.2 Distribution of TTP samples by week

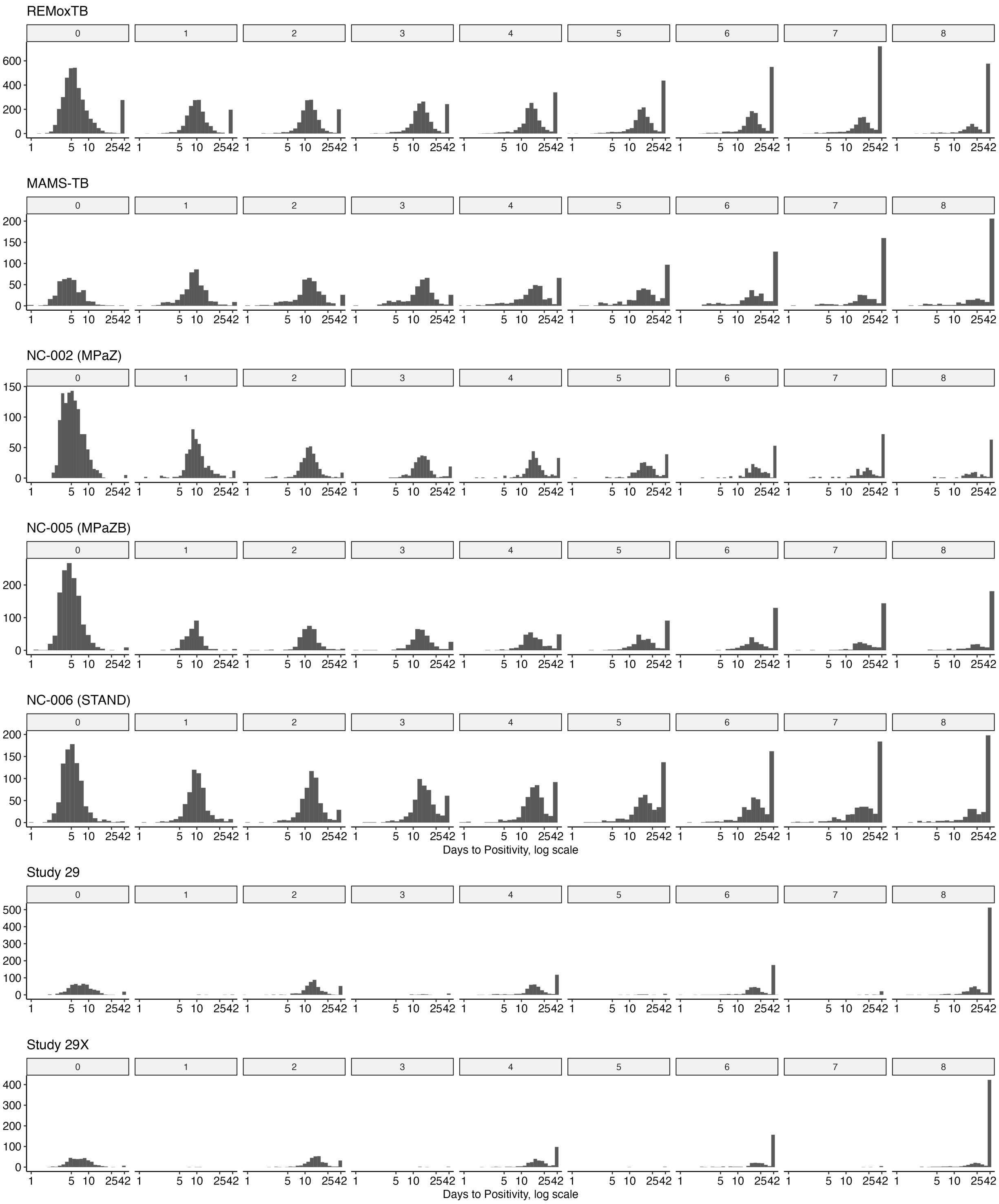


**Fig S1.** Histograms showing the distribution of TTP values for each week's sample. "Negative" values are recorded as 42 days.

### S1.3 Bayesian Model Specification

This section aims to follow best practices in reporting Bayesian analyses by following the Bayesian Analysis Reporting Guidelines [17].

#### S1.3.1 Data variables

The dependent variable ($y_{ijk}$) is the $\log_{10}$(TTP) measured from the sputum sample given by individual $i$ on regimen $j$ at visit $k$ and is modeled as a function of time since randomization (in weeks) ($t_{ijk}$). Let $Q_{ijk}$ be an indicator variable that takes the value 1 when $y_{ijk} \geq \log(\text{ULOQ}_M)$, and 0 otherwise. In other words, $Q_{ijk}$ denotes when a value is above the limit of quantification and therefore cannot be included quantitatively in the model. Instead, these values are handled as "right-censored" [18]. The linear mixed effects model is specified as in Eq. 2, with "brms" default priors used for all parameters except $\gamma_0$.

$$
\begin{aligned}
y_{ijk} &\sim \begin{cases} N(\mu, \sigma_1) & Q_{ijk} = 0 \\ 1 - \Phi(\frac{\text{ULOQ}-\mu}{\sigma_1}) & Q_{ijk} = 1 \end{cases} \\
\mu &= \gamma_{0ij} + \gamma_{1ij} t_{ijk} \\
\sigma_1 &\sim \text{Student } t(3, 0, 2.5) \\
\gamma_{0ij} &\sim N(\gamma_0, s_1) \\
\gamma_{1ij} &\sim N(\gamma_1, s_2) \\
\gamma_0 &\sim N(0, 4^2) \\
\gamma_1 &\sim \text{Flat prior (“brms” default)} \\
s_1 &\sim \text{Student } t(3, 0, 2.5) \\
s_2 &\sim \text{Student } t(3, 0, 2.5)
\end{aligned} \tag{2}
$$

#### S1.3.2 Code and software

All analyses were performed using the "brms" package in the R statistical software. All code used to perform these analyses is available at a GitHub repository managed by the first author (`https://github.com/sdufault15/ttp-lod`).

#### S1.3.3 Model results and fit

Model results are included in S2 at the regimen-level. Model statistics reflecting fit and convergence at the population-level are included in Tables S3 - S9. Figure S3 contains the posterior predictive checks for the main analysis models.

**Table S2.** Posterior point estimates (mode) and 95% highest-density credible intervals (HCI) for the regimen-level slopes from the linear models when the TTP diagnostic LOD and different $ULOQ_M$ thresholds are applied.

| | Diagnostic LOD | $ULOQ_M$ | |
|---|---|---|---|
| Regimen $j$ | 42 Day: $\hat{\gamma}_{1j}$ (95% HCI) | 30 Day: $\hat{\gamma}_{1j}$ (95% HCI) | 25 Day: $\hat{\gamma}_{1j}$ (95% HCI) |
| **PanACEA MAMS-TB** | | | |
| HR20ZM | 0.138 (0.124, 0.154) | 0.132 (0.117, 0.146) | 0.127 (0.114, 0.142) |
| HR20ZQ | 0.121 (0.104, 0.137) | 0.116 (0.101, 0.130) | 0.117 (0.098, 0.129) |
| HR35ZE | 0.144 (0.126, 0.160) | 0.138 (0.123, 0.155) | 0.137 (0.121, 0.155) |
| HRZE | 0.128 (0.116, 0.138) | 0.122 (0.109, 0.131) | 0.120 (0.109, 0.131) |
| HRZQ | 0.122 (0.104, 0.137) | 0.115 (0.098, 0.129) | 0.115 (0.097, 0.128) |
| **REMoxTB** | | | |
| HRZE | 0.095 (0.090, 0.100) | 0.089 (0.085, 0.093) | 0.086 (0.082, 0.090) |
| MHRZ | 0.104 (0.100, 0.109) | 0.097 (0.094, 0.102) | 0.095 (0.091, 0.099) |
| EMRZ | 0.107 (0.102, 0.111) | 0.099 (0.095, 0.103) | 0.097 (0.093, 0.100) |
| **NC-002 (PaMZ)** | | | |
| $Pa_{100}MZ$ | 0.154 (0.137, 0.172) | 0.148 (0.131, 0.167) | 0.149 (0.131, 0.166) |
| $Pa_{200}MZ$ | 0.144 (0.128, 0.158) | 0.138 (0.123, 0.152) | 0.137 (0.123, 0.152) |
| HRZE | 0.126 (0.111, 0.146) | 0.122 (0.109, 0.139) | 0.125 (0.109, 0.141) |
| **NC-005 (BPaMZ)** | | | |
| HRZE | 0.123 (0.111, 0.137) | 0.118 (0.108, 0.130) | 0.119 (0.107, 0.130) |
| $B_{200}PaZ$ | 0.143 (0.130, 0.156) | 0.134 (0.123, 0.146) | 0.132 (0.121, 0.144) |
| $B_{load}PaZ$ | 0.137 (0.127, 0.151) | 0.131 (0.120, 0.142) | 0.128 (0.117, 0.138) |
| **NC-006 (STAND, PaMZ)** | | | |
| HRZE | 0.100 (0.090, 0.108) | 0.094 (0.085, 0.103) | 0.093 (0.085, 0.102) |
| $Pa_{100}MZ$ | 0.104 (0.097, 0.114) | 0.099 (0.091, 0.107) | 0.097 (0.089, 0.105) |
| $Pa_{200}MZ$ | 0.117 (0.107, 0.125) | 0.109 (0.099, 0.118) | 0.106 (0.096, 0.116) |
| $Pa_{200}MZ$ | 0.105 (0.097, 0.113) | 0.099 (0.091, 0.107) | 0.097 (0.088, 0.105) |
| **Study 29** | | | |
| $P_{10}HZE$ | 0.108 (0.101, 0.116) | 0.095 (0.089, 0.101) | 0.091 (0.084, 0.096) |
| HRZE | 0.111 (0.105, 0.119) | 0.099 (0.093, 0.105) | 0.094 (0.088, 0.100) |
| **Study 29X** | | | |
| HRZE | 0.122 (0.107, 0.138) | 0.111 (0.097, 0.125) | 0.110 (0.094, 0.123) |
| $P_{10}HZE$ | 0.134 (0.121, 0.147) | 0.122 (0.111, 0.135) | 0.121 (0.109, 0.133) |
| $P_{15}HZE$ | 0.133 (0.120, 0.148) | 0.121 (0.109, 0.135) | 0.120 (0.107, 0.133) |
| $P_{20}HZE$ | 0.141 (0.125, 0.156) | 0.125 (0.114, 0.141) | 0.126 (0.114, 0.143) |

**Table S3.** REMoxTB model results.

| Coefficient | Estimate | Est.Error | 95% HCI | Rhat | Bulk ESS | Tail ESS |
|---|---|---|---|---|---|---|
| **$ULOQ_M = 25$** | | | | | | |
| $\gamma_0$ | 0.86 | 0.04 | (0.81,0.92) | 1.001 | 2540.6 | 1986.2 |
| $\gamma_1$ | 0.09 | 0.02 | (0.03,0.13) | 1.006 | 550.3 | 246.8 |
| $\sigma_1$ | 0.22 | 0.00 | (0.21,0.22) | 1.002 | 4622.1 | 4396.9 |
| **$ULOQ_M = 30$** | | | | | | |
| $\gamma_0$ | 0.87 | 0.05 | (0.79,0.93) | 1.001 | 2668.6 | 1896.3 |
| $\gamma_1$ | 0.10 | 0.02 | (0.05,0.14) | 1.003 | 1761.4 | 1335.9 |
| $\sigma_1$ | 0.23 | 0.00 | (0.23,0.24) | 1.002 | 4476.4 | 3651.7 |
| **LOD = 42** | | | | | | |
| $\gamma_0$ | 0.87 | 0.08 | (0.77,1.01) | 1.021 | 314.1 | 253.6 |
| $\gamma_1$ | 0.10 | 0.03 | (0.06,0.16) | 1.005 | 574.3 | 508.0 |
| $\sigma_1$ | 0.25 | 0.00 | (0.25,0.26) | 1.005 | 974.4 | 1751.1 |

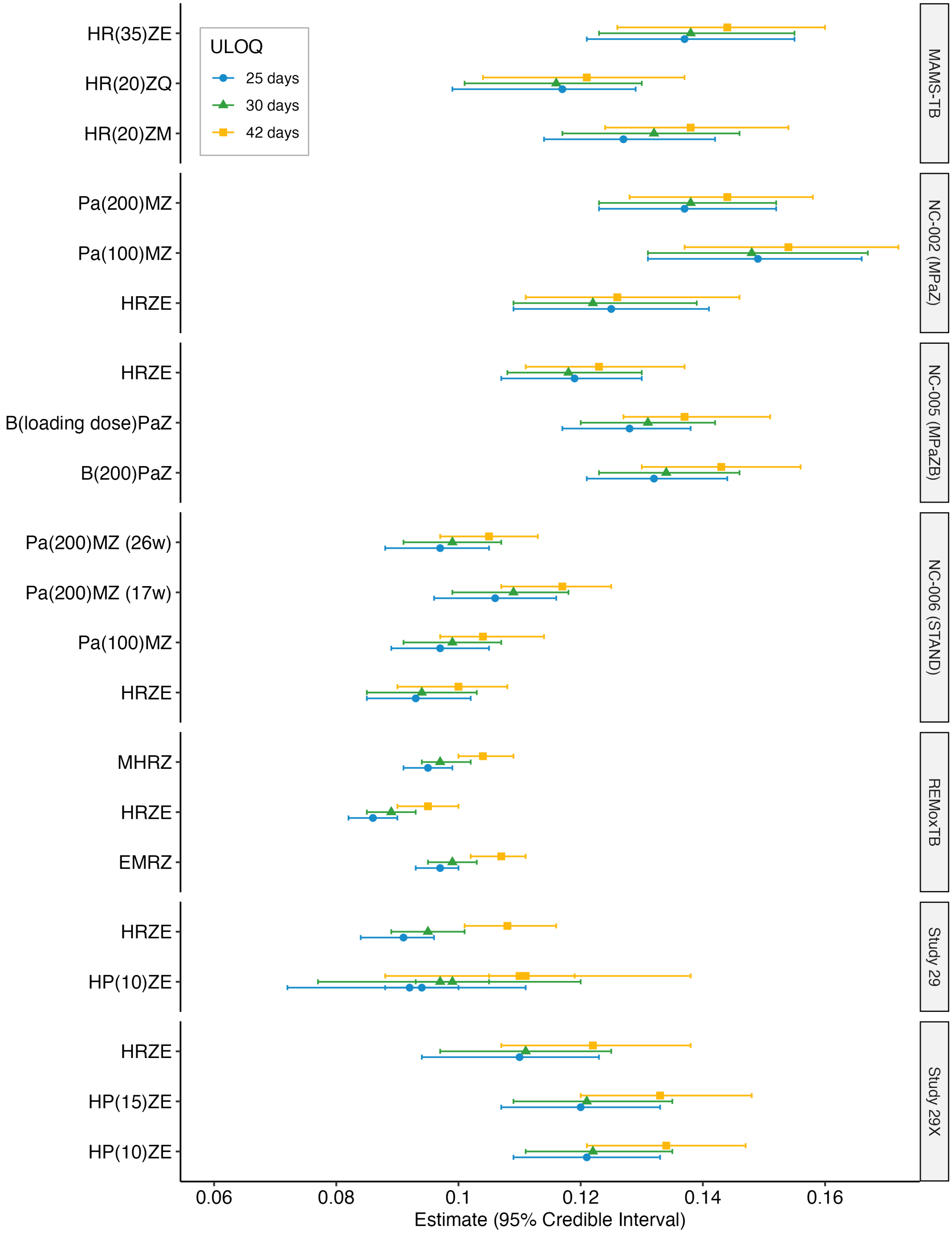


**Fig S2.** Forest plot of regimen-level estimates and 95% HCI

**Table S4.** PanACEA MAMS-TB model results.

| Coefficient | Estimate | Est.Error | 95% HCI | Rhat | Bulk ESS | TailESS |
|---|---|---|---|---|---|---|
| **ULOQ$_M$ = 25** | | | | | | |
| $\gamma_0$ | 0.76 | 0.02 | (0.73,0.80) | 1.000 | 2954.7 | 2264.4 |
| $\gamma_1$ | 0.12 | 0.01 | (0.10,0.15) | 1.005 | 1100.1 | 751.3 |
| $\sigma_1$ | 0.26 | 0.00 | (0.25,0.27) | 1.000 | 2542.7 | 2291.0 |
| **ULOQ$_M$ = 30** | | | | | | |
| $\gamma_0$ | 0.76 | 0.02 | (0.73,0.80) | 1.000 | 3045.1 | 2320.7 |
| $\gamma_1$ | 0.12 | 0.01 | (0.10,0.15) | 1.004 | 1433.4 | 1001.3 |
| $\sigma_1$ | 0.26 | 0.00 | (0.26,0.27) | 1.002 | 2983.7 | 2721.8 |
| **LOD = 42** | | | | | | |
| $\gamma_0$ | 0.76 | 0.02 | (0.72,0.80) | 1.001 | 3390.1 | 2943.4 |
| $\gamma_1$ | 0.13 | 0.01 | (0.11,0.16) | 1.002 | 1527.9 | 1009.2 |
| $\sigma_1$ | 0.28 | 0.00 | (0.27,0.28) | 1.001 | 3168.5 | 2510.8 |

**Table S5.** NC-002 (PaMZ) model results.

| Coefficient | Estimate | Est.Error | 95% HCI | Rhat | Bulk ESS | Tail ESS |
|---|---|---|---|---|---|---|
| **ULOQ$_M$ = 25** | | | | | | |
| $\gamma_0$ | 0.75 | 0.08 | ( 0.62,0.90) | 1.006 | 702.6 | 1011.4 |
| $\gamma_1$ | 0.13 | 0.09 | (-0.04,0.26) | 1.028 | 179.9 | 123.0 |
| $\sigma_1$ | 0.15 | 0.00 | ( 0.14,0.15) | 1.005 | 623.2 | 1262.0 |
| **ULOQ$_M$ = 30** | | | | | | |
| $\gamma_0$ | 0.75 | 0.08 | ( 0.61,0.89) | 1.003 | 752.5 | 887.7 |
| $\gamma_1$ | 0.14 | 0.08 | ( 0.04,0.33) | 1.017 | 377.5 | 170.9 |
| $\sigma_1$ | 0.15 | 0.00 | ( 0.15,0.16) | 1.002 | 1168.0 | 2327.6 |
| **LOD = 42** | | | | | | |
| $\gamma_0$ | 0.75 | 0.11 | ( 0.57,0.92) | 1.047 | 275.1 | 546.0 |
| $\gamma_1$ | 0.14 | 0.06 | (-0.01,0.25) | 1.011 | 405.7 | 302.6 |
| $\sigma_1$ | 0.16 | 0.00 | ( 0.16,0.17) | 1.010 | 289.8 | 584.2 |

**Table S6.** NC-005 (BPaMZ) model results.

| Coefficient | Estimate | Est.Error | 95% HCI | Rhat | Bulk ESS | Tail ESS |
|---|---|---|---|---|---|---|
| **ULOQ$_M$ = 25** | | | | | | |
| $\gamma_0$ | 0.73 | 0.05 | (0.66,0.82) | 1.002 | 2053.8 | 2159.1 |
| $\gamma_1$ | 0.13 | 0.04 | (0.06,0.20) | 1.007 | 982.6 | 723.5 |
| $\sigma_1$ | 0.16 | 0.00 | (0.16,0.16) | 1.000 | 3552.5 | 4313.0 |
| **ULOQ$_M$ = 30** | | | | | | |
| $\gamma_0$ | 0.73 | 0.07 | (0.65,0.84) | 1.003 | 1030.1 | 1650.8 |
| $\gamma_1$ | 0.13 | 0.04 | (0.04,0.21) | 1.004 | 726.2 | 523.9 |
| $\sigma_1$ | 0.17 | 0.00 | (0.16,0.17) | 1.006 | 1368.0 | 2715.9 |
| **LOD = 42** | | | | | | |
| $\gamma_0$ | 0.72 | 0.07 | (0.61,0.82) | 1.003 | 1259.5 | 1976.4 |
| $\gamma_1$ | 0.13 | 0.05 | (0.03,0.23) | 1.007 | 962.3 | 1082.0 |
| $\sigma_1$ | 0.18 | 0.00 | (0.17,0.18) | 1.002 | 2516.9 | 3924.4 |

**Table S7.** NC-006 (STAND, PaMZ) model results.

| Coefficient | Estimate | Est.Error | 95% HCI | Rhat | Bulk ESS | Tail ESS |
|---|---|---|---|---|---|---|
| **$\mathbf{ULOQ}_M$ = 25** | | | | | | |
| $\gamma_0$ | 0.83 | 0.03 | (0.79,0.89) | 1.002 | 2943.0 | 2333.9 |
| $\gamma_1$ | 0.09 | 0.01 | (0.08,0.10) | 1.001 | 2896.1 | 3108.1 |
| $\sigma_1$ | 0.20 | 0.00 | (0.20,0.21) | 1.001 | 3053.1 | 5209.5 |
| **$\mathbf{ULOQ}_M$ = 30** | | | | | | |
| $\gamma_0$ | 0.84 | 0.02 | (0.78,0.89) | 1.001 | 2769.6 | 3095.5 |
| $\gamma_1$ | 0.08 | 0.01 | (0.07,0.10) | 1.002 | 2234.3 | 1782.1 |
| $\sigma_1$ | 0.20 | 0.00 | (0.20,0.21) | 1.003 | 2629.9 | 3753.3 |
| **LOD = 42** | | | | | | |
| $\gamma_0$ | 0.84 | 0.03 | (0.79,0.90) | 1.004 | 1889.5 | 1674.5 |
| $\gamma_1$ | 0.08 | 0.01 | (0.07,0.09) | 1.002 | 2354.7 | 2054.0 |
| $\sigma_1$ | 0.20 | 0.00 | (0.19,0.21) | 1.006 | 1371.4 | 4309.1 |

**Table S8.** Study 29 model results.

| Coefficient | Estimate | Est.Error | 95% HCI | Rhat | Bulk ESS | Tail ESS |
|---|---|---|---|---|---|---|
| **$\mathbf{ULOQ}_M$ = 25** | | | | | | |
| $\gamma_0$ | 0.92 | 0.07 | (0.80,1.05) | 1.003 | 4367.8 | 3765.9 |
| $\gamma_1$ | 0.09 | 0.02 | (0.06,0.13) | 1.002 | 2919.8 | 617.7 |
| $\sigma_1$ | 0.20 | 0.00 | (0.19,0.21) | 1.001 | 2064.0 | 702.8 |
| **$\mathbf{ULOQ}_M$ = 30** | | | | | | |
| $\gamma_0$ | 0.92 | 0.08 | (0.77,1.08) | 1.002 | 3536.7 | 2537.8 |
| $\gamma_1$ | 0.10 | 0.02 | (0.05,0.14) | 1.006 | 2133.2 | 1573.3 |
| $\sigma_1$ | 0.21 | 0.00 | (0.20,0.22) | 1.000 | 1764.5 | 3103.1 |
| **LOD = 42** | | | | | | |
| $\gamma_0$ | 0.92 | 0.14 | (0.73,1.12) | 1.001 | 2081.2 | 1029.8 |
| $\gamma_1$ | 0.11 | 0.03 | (0.07,0.16) | 1.002 | 2727.5 | 1818.1 |
| $\sigma_1$ | 0.23 | 0.01 | (0.22,0.24) | 1.001 | 1976.1 | 3644.8 |

**Table S9.** Study 29X model results.

| Coefficient | Estimate | Est.Error | 95% HCI | Rhat | Bulk ESS | Tail ESS |
|---|---|---|---|---|---|---|
| **$\mathbf{ULOQ}_M$ = 25** | | | | | | |
| $\gamma_0$ | 0.89 | 0.03 | (0.85,0.94) | 1.001 | 4674.3 | 4053.9 |
| $\gamma_1$ | 0.12 | 0.02 | (0.09,0.15) | 1.005 | 1889.7 | 1803.0 |
| $\sigma_1$ | 0.20 | 0.01 | (0.19,0.21) | 1.002 | 2117.6 | 4125.4 |
| **$\mathbf{ULOQ}_M$ = 30** | | | | | | |
| $\gamma_0$ | 0.90 | 0.03 | (0.85,0.95) | 1.000 | 6261.9 | 5252.2 |
| $\gamma_1$ | 0.12 | 0.01 | (0.09,0.15) | 1.002 | 2041.5 | 1515.1 |
| $\sigma_1$ | 0.20 | 0.01 | (0.19,0.21) | 1.002 | 2375.2 | 4456.4 |
| **LOD = 42** | | | | | | |
| $\gamma_0$ | 0.89 | 0.03 | (0.84,0.94) | 1.001 | 5871.9 | 4704.7 |
| $\gamma_1$ | 0.13 | 0.02 | (0.10,0.16) | 1.000 | 2352.5 | 1539.9 |
| $\sigma_1$ | 0.22 | 0.01 | (0.21,0.23) | 1.001 | 2101.2 | 4780.5 |

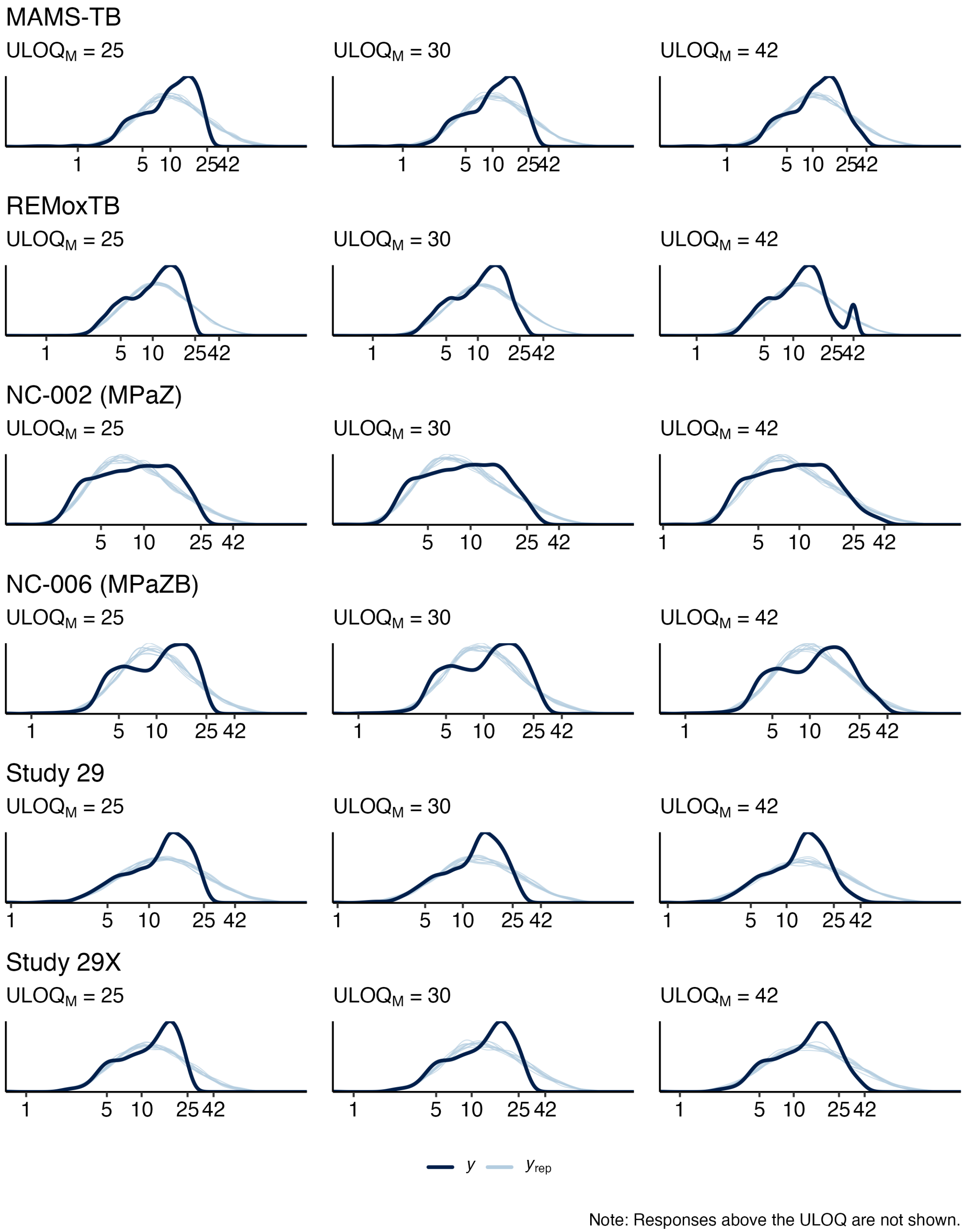


**Fig S3.** Posterior predictive checks comparing the observed response data ($y$) to data drawn from the posterior of the model ($y_{\text{rep}}$).

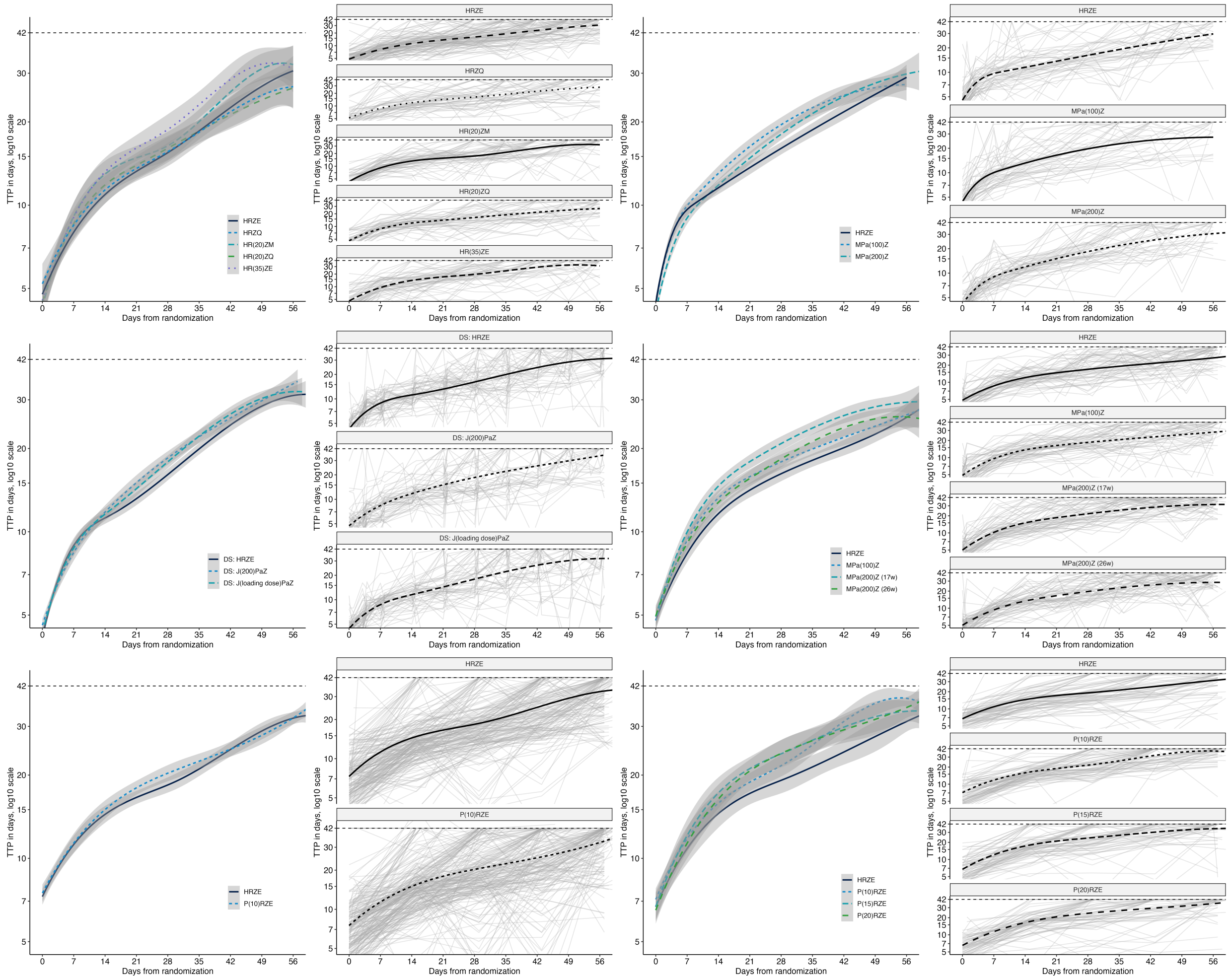


**Fig S4. Observed time-to-positivity trajectories.** Any observations at or above the diagnostic limit of detection (42 days) are recorded as 42 days. A: Regimen-level trends in TTP (lines) and estimated STAND, PaMZard errors (ribbons) as fit by smoothing splines. B: Individual TTP trajectories (light gray) and regimen-level smoothing spline (black).

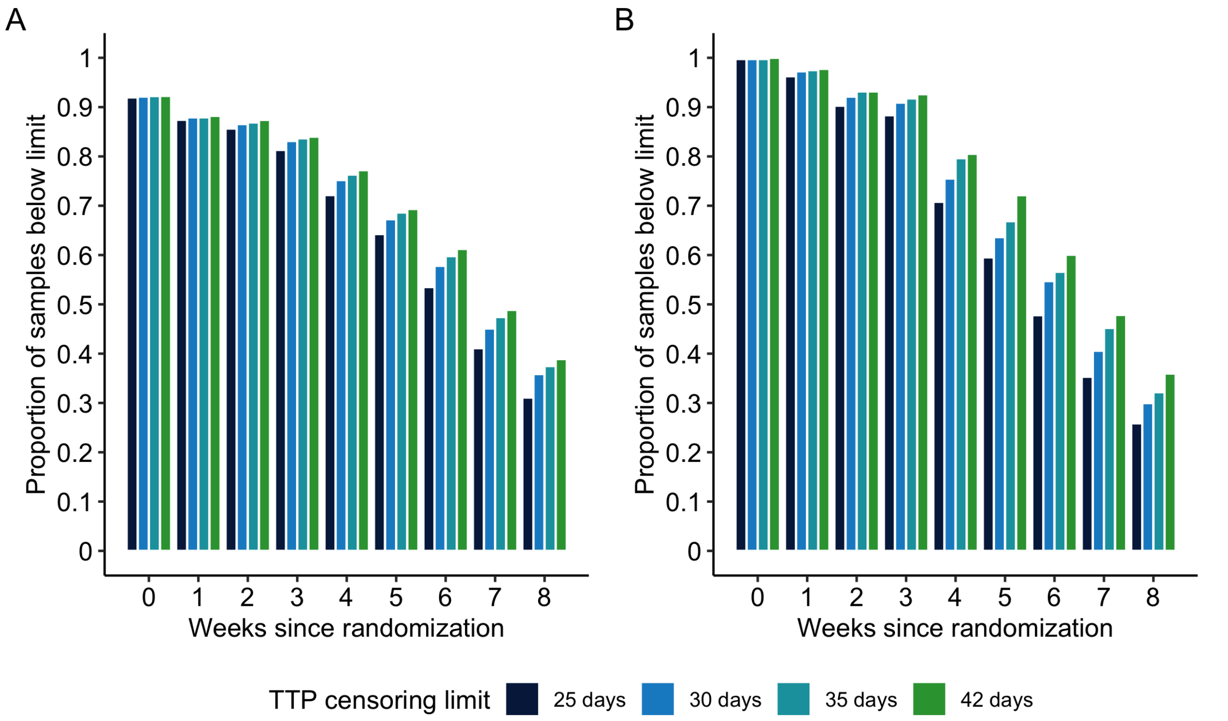


**Fig S5.** The proportion of samples that return TTP observations below the BACTEC MGIT diagnostic LOD (42 days, green) as well as below various $\text{ULOQ}_M$s for each week since randomization for (A) REMox-TB and (B) PanACEA MAMS-TB data.

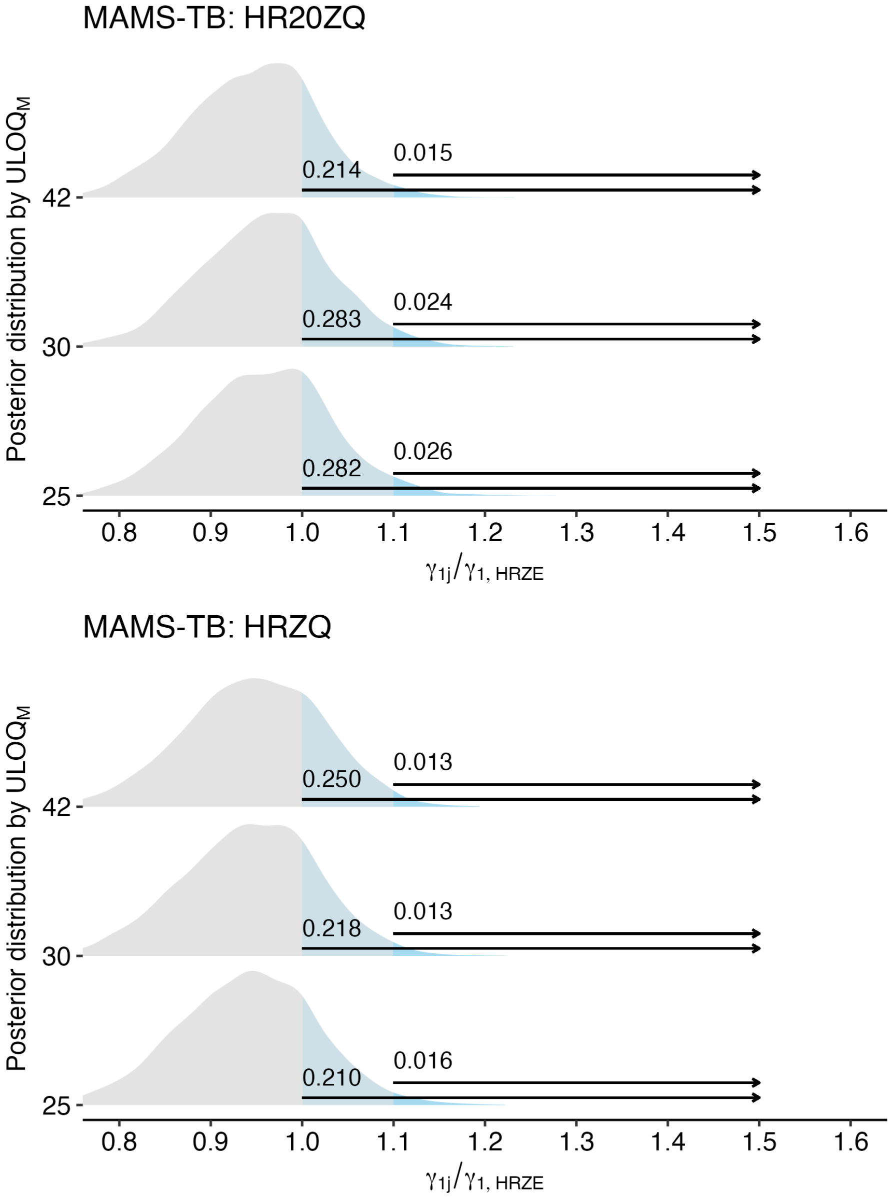


**Fig S6.** Among regimens with similar or worse bactericidal activity as HRZE, the posterior distributions for the relative comparison of a regimen's slope ($\gamma_{1j}$) against the estimated slope on HRZE ($\gamma_{1,\text{HRZE}}$), where a value of 1 indicates equal slopes values ($> 1$) suggest the regimen has greater bactericidal activity than HRZE. The estimated "confidence" that a regimen has any improvement in bactericidal activity over HRZE ($\Pr(\gamma_{1j}/\gamma_{1,\text{HRZE}} > 1)$) as well as the "confidence" that a regimen has more than 10% improvement in bactericidal activity over HRZE ($\Pr(\gamma_{1j}/\gamma_{1,\text{HRZE}} > 1.1)$) is indicated for each regimen at each ULOQ$_M$. Annotated are the corresponding values for the posterior probabilities of $\Pr(\gamma_{1j}/\gamma_{1,\text{HRZE}} > \tau)$, where $\tau$ equals 1 and 1.1, respectively.
